# Biallelic *MCUR1* nonsense mutation associated with vacuolar myopathy and altered mitochondrial calcium signaling

**DOI:** 10.1101/2025.10.17.25338070

**Authors:** Anna Maria Haschke, Anja von Renesse, Eugenio Graceffo, Susanne Morales-Gonzalez, Alessandro Prigione, Christoph Hübner, Werner Stenzel, Markus Schuelke

## Abstract

During muscle contraction, increased influx of mitochondrial calcium (mtCa²⁺) from the myocyte cytosol through the mitochondrial calcium uniporter (MCU) couples calcium homeostasis with high ATP provision. The mitochondrial calcium uniporter regulator 1 (MCUR1) is an integral membrane protein that promotes MCU activity. Although its function has been studied in cell models, mutations in MCUR1 have not yet been associated with human disease. Here, we present a case study of a patient exhibiting proximal muscle weakness and atrophy, who carries a novel homozygous loss-of-function mutation in MCUR1. To investigate the underlying mechanisms of muscle pathology, we examined patient fibroblasts and quadriceps muscle specimens. MCUR1 deficiency compromised mitochondrial Ca²⁺ uptake upon histamine exposure, but did not alter resting mitochondrial membrane potential or MCU protein complex assembly or subcellular location. Consequently, ATP production and oxygen consumption were reduced, and mitochondrial biogenesis was disturbed in muscle, with histological features of autophagic vacuoles with sarcolemmal features. Our study associates MCUR1 deficiency with mitochondrial dysfunction and autophagic vacuolar myopathy, thereby highlighting the crucial role of mitochondrial Ca²⁺ uptake in regulating mitochondrial function and expanding the spectrum of mitochondrial disorders in humans.

**Graphical abstract:** 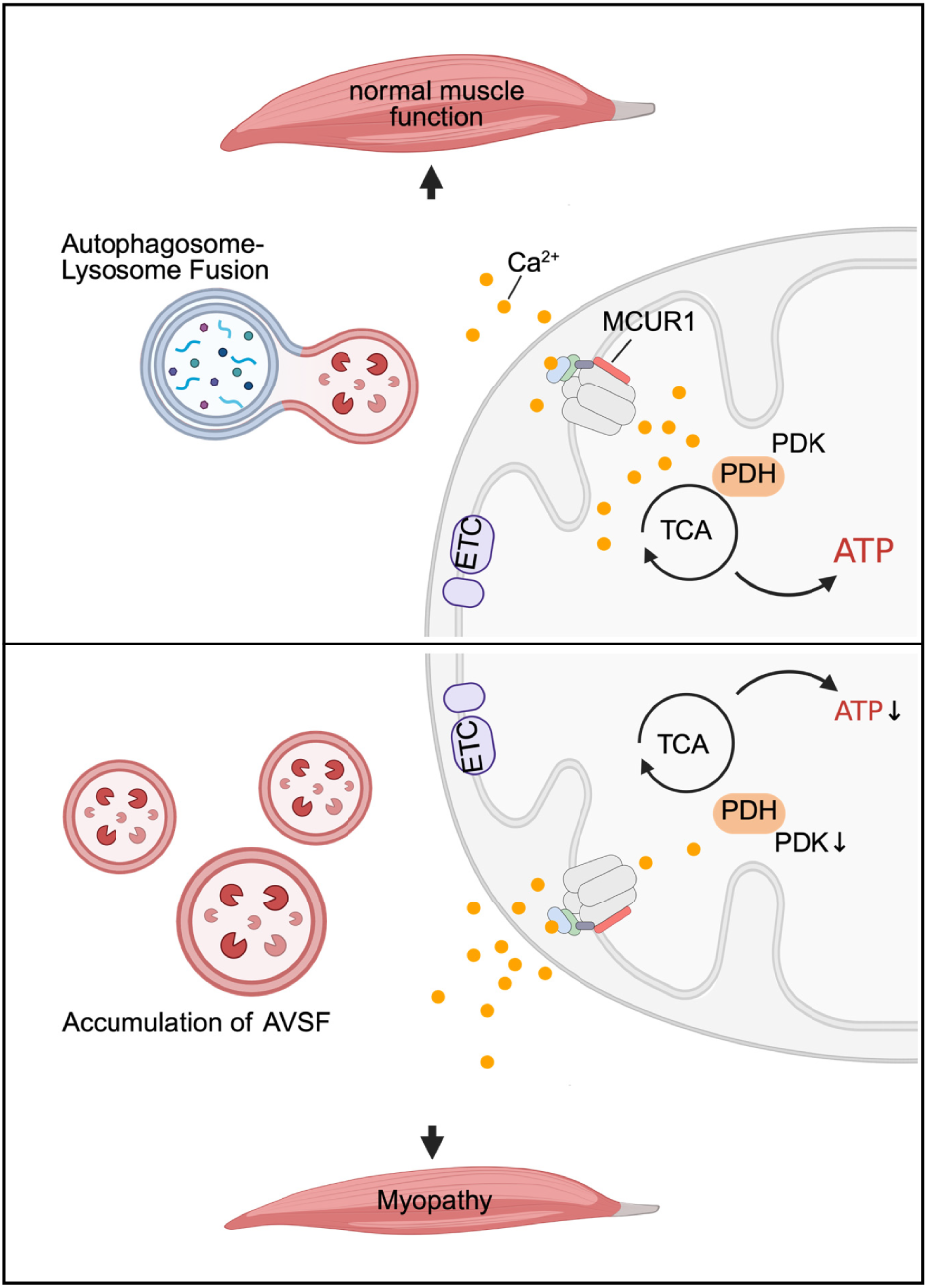

## Introduction

Muscle contraction is a highly orchestrated process, by which calcium transients are released from the sarcoplasmic reticulum (SR) into the cytosol of the myocyte to activate the contractile elements. Mitochondria play a crucial role in regulating these calcium transients, mainly by removing the cytosolic Ca^2+^-ions and pumping them back into the SR.^1^ This process is closely coupled with the oxidative phosphorylation that generates the negative mitochondrial membrane potential (MMP) and allows ATP production by the F_1_F_0_-ATPase. The highly negative voltage gradient across the inner mitochondrial membrane (IMM), known as the mitochondrial membrane potential (ΔΨ_M_), drives the mtCa^2+^ uptake. Ca²⁺ enters into the mitochondrial matrix *via* the mitochondrial calcium uniporter (MCU). Under physiological conditions, calcium buffering through the mitochondria is central for ATP production, redox balance, and regulation of autophagy.^1^ The mtCa^2+^ uniporter complex consists of three core pore-forming proteins [associated *gene* names], MCU [*MCU*], MCUb [*MCUB*], and EMRE [*SMDT1*] and of additional regulatory subunits (MICU1 [*MICU1*], MICU2 [*MICU2*], MICU3 [*MICU3*], and MCUR1 [*MCUR1*]).^2–4^

In 2013, Pan *et al.* generated an *MCU*^−/−^ mouse model by placing a gene trap into *MCU* intron 1. Homozygous animals had a reduced body size and exhibited exercise intolerance. Isolated muscle mitochondria failed to take up calcium from the surrounding medium.^5^ Further studies silenced the *MCU* gene with short hairpin RNAs (shRNA) against *MCU* that had been delivered by adeno-associated virus serotype 9 (AAV9) and confirmed these initial findings. *MCU* knock-down animals had muscle atrophy, distorted mitochondrial shapes, and an increased number of vacuoles. In contrast, *MCU* overexpression *via* AAV9-mediated gene transfer led to muscle hypertrophy.^6,7^ At present, no patients have been described, who carry mutations in the coding sequences of the MCU pore-forming proteins.

However, researchers discovered recessive mutations in the MICU1 regulatory subunit. Patients suffered from early-onset proximal muscle weakness and intellectual disability.^8,9^ Another regulatory component of the MCU complex is the mitochondrial calcium uniporter regulator 1 (MCUR1), which is an integral membrane protein of the IMM. MCUR1 acts as scaffold protein and promotes MCU activity. Its binding to both MCU and EMRE facilitates the assembly and stability of the MCU complex. Abrogation of MCUR1 in mouse cardiomyocytes and in endothelial cells disrupts the oligomerization of the MCU complex, underlining its importance in forming an active MCU complex.^10^ Functional studies in HEK293T and HeLa cells have shown that *MCUR1* knock-down by RNA interference (RNAi) resulted in decreased mitochondrial Ca²⁺ uptake and basal oxygen consumption, as well as impaired oxidative phosphorylation (OXPHOS) and ATP production. This confirms the effects observed in *MCU* knockout models.^11,12^ These findings support the hypothesis that MCUR1 is critical for MCU-mediated Ca²⁺ uptake.

So far, *MCUR1* mutations have not yet been associated with human genetic disease. Here we present the case of a patient with a homozygous recessive nonsense mutation in *MCUR1*, and describe his muscle histology, calcium signaling, and effects on downstream regulatory pathways that are essential for maintaining muscle health.

## Materials and methods

### Consent

Written informed consent was obtained from the family of the patient. The study was approved by the institutional review board of Charité (EA2/202/08). All participants gave written informed consent in accordance with the Declaration of Helsinki for all aspects of the study. Anonymised control fibroblast lines and muscle biopsy specimens were obtained from leftover diagnostic samples from patients without neuromuscular disorders.

### Statistical analysis

We used Graph Pad Prism v10.4.1 software to run all statistical analyses, which are detailed in the figure captions. Data are presented as mean ± standard deviation (SD), unless specified otherwise. Details on the statistical tests can be found in the corresponding figure legends.

### Fibroblast cell culture

Fibroblasts were derived from skin punch biopsy samples and cultured in DMEM (Gibco, #41966-029) supplemented with 15% foetal bovine serum (Gibco, #10500-064) and 1% penicillin–streptomycin (Gibco, #15140122), and incubated at 37°C in 5% CO_2_. The fibroblast line was confirmed to harbor the correct homozygous *MCUR1* mutation by Sanger sequencing prior to further functional studies. Cell lines were routinely tested for mycoplasma contamination.

### DNA analysis

Whole exome sequencing was performed using a blood DNA sample of the index patient. The coding exons and flanking intronic regions were enriched using the Agilent SureSelect XT Human All Exon V5 Exome Kit and sequenced as 100 bp paired-end fragments on an Illumina HiSeq2500 machine. The 65 million resulting sequence fragments (FASTQ files) were aligned to the human reference genome (GRCh37.p11, hg19/Ensembl 72) using the BWA-MEM v0.7.8 software. Deviations from the reference sequence were detected and recorded using the GATK v3.1.1 software package and exported as a VCF-file. Each of these variants was then analysed using the MutationTaster2^13^ variant assessment software to identify potentially pathogenic variants. Segregation analysis of the variants in the family was performed by Sanger sequencing. Oligonucleotide primers (see Supplementary Table 1) were used to amplify exon 6 of *MCUR1* and sequencing was performed with the dideoxy chain termination method (BigDye Terminator Version 3.1, Thermo Fisher, #4337457) on an ABI 3730 DNA sequencer.

### RNA analysis

For RNA-sequencing, total RNA was isolated from patient and control muscle biopsy samples that had been obtained from the quadriceps muscle and from cultured skin fibroblast specimens using using the TRIzol protocol (Invitrogen, #15596026). 500 ng of RNA from each sample were sent for bulk RNA sequencing to the Beijing Genomics Institute on their DNBseq-500 platform. The library preparation included an mRNA enrichment step using oligo(dT)-attached magnetic beads. The strand-specific cDNA library generated 60 million 100 bp paired-end reads. Sequence quality was then assessed using FastQC v0.11.8 and MultiQC v1.6.^14^ Reads were mapped to the EMBL human genome 38, patch release 13, using the splice-aware aligner STAR v2.7.10a.^15^ BAM files were sorted and indexed using SAMtools v1.9.^16^ StringTie v2.1.7 was used to generate the gene count matrix.^17^ Normalization of fragment counts, PCA, clustering and differential expression analysis were performed in R v4.3.2 with the DESeq2 v1.42.0 package. For visualizing purposes, LFC shrinkage using “apeglm” was performed.^18^ Unless stated otherwise, an FDR<0.1 was considered statistically significant in DESeq2 analysis. GO enrichment Analysis of Biological Processes was performed using ShinyGO v0.741.^19^

### qPCR analysis

We performed transcript level analysis using quantitative real time-PCR. Total mRNA was converted to cDNA using the SuperScript III First-Strand cDNA system (Thermo Fisher, #18091050). PCR analysis of cDNA was performed using primers specific for human *MCUR1* and the MCU complex subunit genes (see Supplementary Table 1). Each reaction was run in triplicate. Amplimer amounts were quantified continuously with the SYBR Green qPCR system (Applied Biosystems, #A46012) using an qTower3 (Analytik Jena). The *GAPDH* signal was used for normalization.

### Immunofluorescence staining

Fibroblast cells were seeded onto glass coverslips and cultivated at 37°C and 5% CO_2_ until they reached 80% confluence. The mitochondria were stained with 250 nM MitoTracker Red (Thermo Fisher, M7512) in Opti-MEM (Reduced Serum Medium, Thermo Fisher, 31985-062) for 30 min under growth conditions. The cells were then fixed with a 4% PFA solution in PBS for 15 min and rinsed three times with PBS. To permeabilize the cells, they were incubated for 10 min with 0.1% Triton X-100 (Merck, 108643) in PBS under continuous rocking. After blocking with 10% normal goat serum (Abcam, ab7475) in PBS for 1 h, the cells were incubated with a monoclonal goat anti-MCU antibody (D2Z3B clone, dilution 1:100 dilution, Cell Signaling, #14997) overnight at 4°C. After washing, the cells were incubated with the secondary antibody (goat anti-rabbit Alexa Fluor 488, Thermo Fisher A3273, dilution 1:200 with 10% normal donkey serum) for 45 min at RT under light protection. Counterstaining of the nuclei was done with DAPI (dilution 1:10,000; Invitrogen, D1306). Cells were mounted on Mowiol 4-88 Mounting Media (ROTH, 0713). After immunostaining, cells were imaged with a Leica Thunder DMi8 microscope.

### Western blotting

Cells were lysed in RIPA lysis buffer [1% NP-40, 0.1% SDS, 50 mM Tris-HCl pH 8, 150 mM NaCl, 0.5% sodium deoxycholate, and Complete^TM^ EDTA-free protease inhibitor (Roche, 11 873 580 001)] and quantified with the Protein Assay Dye Reagent Concentrate (Bio-Rad, #5000001). Protein was separated on a NuPAGE 4-12%, Bis-Tris gel (Thermo Fisher, NP0329), transferred to nitrocellulose membrane (Amersham, #10600001) and stained with Ponceau solution. Protein bands were visualized using specific antibodies (see **Supplementary Table 1**).

### Histology and histochemistry

All histological and histochemical studies were performed on cryosections of the quadriceps muscle. Histological stainings were imaged with a Leica DMLB microscope. For morphometric analysis, the muscle fibers’ Ferret diameter was determined with the ImageJ v1.45 software.

### Functional assessment of patient material

#### Oxygen consumption rate

The oxygen consumption rate (OCR) was measured with a XF HS Mini Analyzer (Agilent) and XF Cell Mito Stress Test Kit (Agilent, #103010). 10,000 cells/well were seeded in a 8-well Seahorse XF Cell Culture Microplate (Agilent, #103725) and incubated overnight at 37°C. Prior to the assay, growth media was replaced with XF DMEM medium, pH 7.4 (Agilent, #103575) supplemented with 1 mM sodium pyruvate (Sigma-Aldrich, #S8636) 2 mM L-glutamine (Gibco, #25030149) and 10 mM glucose (Sigma-Aldrich, #49163). Cells were then incubated for 45-60 min in 37°C without CO_2_ to prevent acidification of the medium. The basal respiration rate was measured before cells were exposed sequentially to oligomycin (1 μM), carbonyl cyanide p-trifluoromethoxyphenylhydrazone (FCCP 1.5 μM) and rotenone + antimycin A (500 nM each). After each injection, the OCR was measured for 5 min, the medium was mixed and again measured for another 5 min. After the experiment, the cell number was determined by CyQUANT (Invitrogen, #C35011) for normalization.

#### Assessment of live-cell ATP production

Fibroblasts were detached by 0.05% Trypsin-EDTA (Gibco, #25300054) treatment and pcDNA3.2-V5-mtLuc (Addgene, #219666) was introduced using Amaxa Nucleofector 2 (Lonza) and the Amaxa Human Dermal Fibroblast Nucleofector Kit (Lonza, VPD-1001) according to protocol. This vector targets the firefly luciferase into the mitochondrial matrix. In the presence of luciferin, the mitochondria emit light whose intensity is proportional to the ATP concentration in the mitochondrial matrix. After addition of 1 μM cell permeable DMNPE-caged Luciferin (Abcam, ab145163) we measured bioluminescence using the GloMax plate reader (Promega, GM3000). Data was normalized to the number of cells evaluated by CyQUANT (Invitrogen, #C35011).

#### Intracellular Ca^2+^ imaging

The cells were transfected using Nucleofector as described before using the plasmids pCMV_R-GECO1 (Addgene #32444, measures intra-cytosolic Ca^2+^, red fluorescence) and pCMV_CEPIA2mt (Addgene #58218, measures intra-mitochondrial Ca^2+^, green fluorescence). For all live cell experiments, the cells were seeded in a m-Dish 35 mm with glass bottom (ibidi, 81158), mounted on a microscope stage and continuously perfused using a peristaltic perfusion pump (PPS5, Multichannel systems) at the speed of 1 ml*min^−1^ with physiological salt solution (PSS) containing [mM] 150 NaCl, 4 KCl, 2 CaCl_2_, 1 MgCl_2_, 5.6 glucose and 25 HEPES (pH 7.4). After an interval of 10 μM histamine was spiked into the perfusion solution. Time-lapse images of fluorescence intensity were captured with a Leica DMi8 using a 20×, 1.4 NA lens at 37°C in 5% CO_2_. The intra-mitochondrial Ca^2+^ was recorded with a [green, Ex 488 nm | Em 510 nm] and the cytosolic Ca^2+^ was simultaneously recorded with a [red, Ex 555 nm | Em 584 nm] setting. Images were taken at a rate of one frame per second.

#### Ψm measurement in living cells

Cells were equilibrated with Tetramethylrhodamine methyl ester (TMRM, 25 nM) and Hoechst 33347 (50 nM) for 30 min at 37°C, 5% CO_2_. Images were acquired on Leica DMi8 using a 20×, 1.4 NA lens. Fluorescence was excited at 535 nm and measured at 584 nm. Cell profiler analysis software was used for image analysis.^20^

#### Determination of the mtDNA copy number

Relative mtDNA copy numbers were determined by qPCR using the amplification curves of a gene encoded on the mtDNA (*MT-ND1*, NC_012920.1) and a single copy gene on the nuclear DNA (*NDUFV1,* NM_001166102.2) using oligonucleotide primers (see Supplemental table 1) with the SYBR green technology on a TRIO combi cycler (Biometra, 846-2-070-720).

## Results

### MCUR1 loss-of-function is associated with proximal muscle weakness and muscle atrophy

We describe an adolescent patient, son of first degree cousins with two healthy sisters, who experienced mild proximal muscle weakness and atrophy in his legs from from his early school years onward, later progressing to his lower thighs and feet (progressive cavus foot deformity) and hands (interdigital muscle atrophy, **Figure 1A**). In his early teen years, his muscle weakness had progressed and he showed a positive Gowers sign. His creatine kinase (CK) levels were constantly elevated to >3,800 U/L (N<190) suggestive of myofiber damage and fibrosis, which could be confirmed by muscle ultrasound. Serum lactate levels were normal [14.6 mg/dl, N<19], but his pyruvate levels were mildly increased [1.5 mg/dl, N<0.9]. EMG confirmed myopathy with massive spontaneous activity, polyphasic potentials, and reduced motor unit potential amplitudes. Motor and sensor neurography, visual evoked potentials (VEP), EEG, and intellectual function were all normal. He did not have a cardiomyopathy, but Wolff-Parkinson-White syndrome was successfully treated by ablation in his late teen years. The histological investigation of his muscle biopsy specimen has been reported in detail before^21^ and revealed an autophagic vacuolar myopathy with variation in fiber size, and increased frequency of accumulated vacuoles (**Figure 1C, Supplementary Figure 1A**).

**Figure 1.**
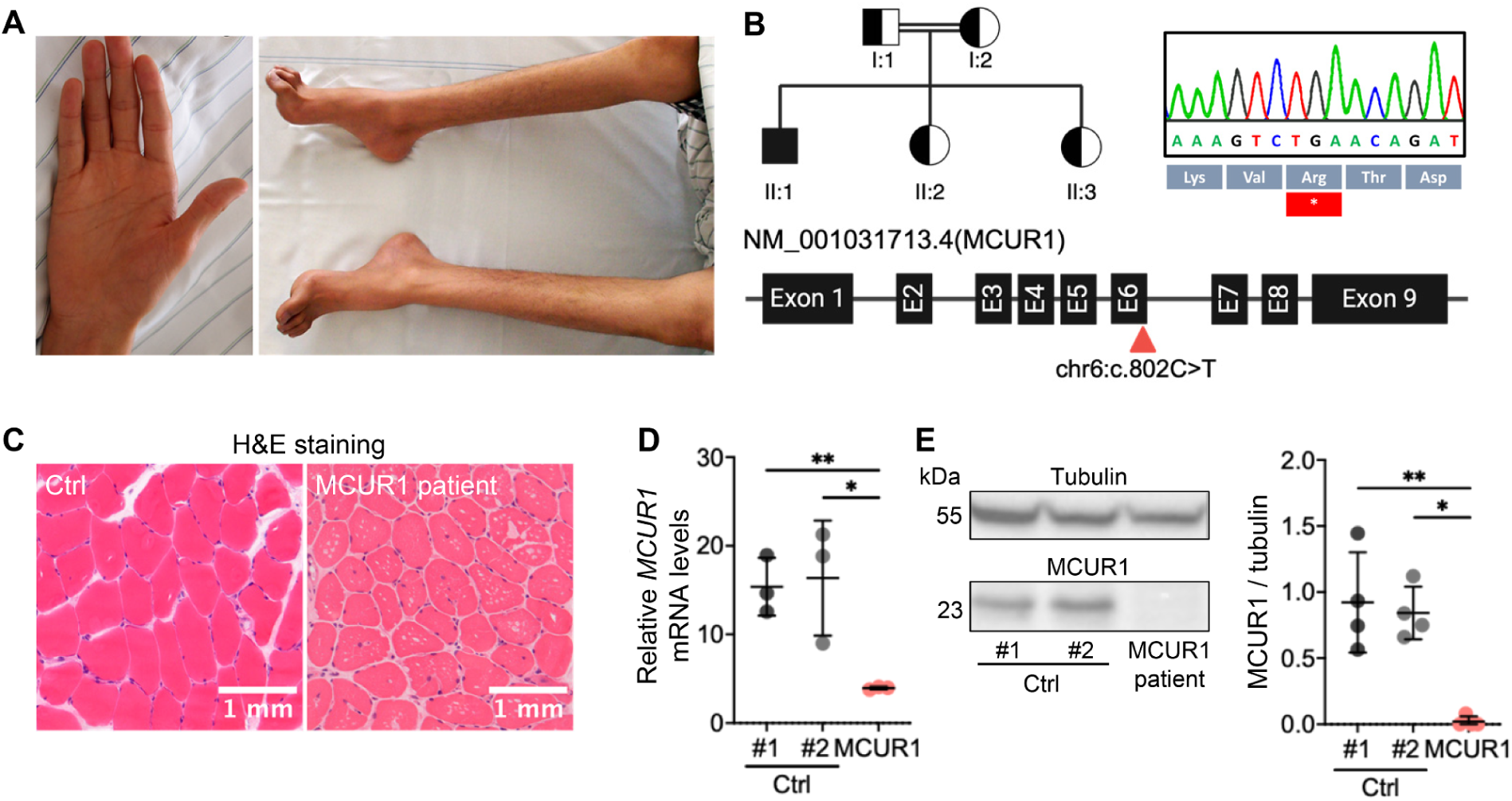
Loss-of-function mutations in *MCUR1* identified in a patient with muscle atrophy. **(A)** Patient’s hands presenting thenar muscle atrophy and feet with a high arch (cavus deformity). **(B)** Pedigree of the consanguineous family with one affected child (filled symbol). The *MCUR1* variant in exon 6 was confirmed by Sanger sequencing and annotated on the *MCUR1* transcript (NM_006077.3). **(C)** Hematoxylin and eosin staining of healthy and patient-derived quadriceps biopsies showing central nuclei and multiple vacuoles inside the muscle fibers. **(D)** Scatter plot of the qPCR analysis demonstrating a significant reduction of *MCUR1* mRNA copy numbers in the MCUR1-deficient patient fibroblasts [*MCUR1*/1,000 *GAPDH* copies]; error bars represent the mean ± standard deviation (SD) of three biological replicates. Expression was normalized to GAPDH. **(E)** Immunoblot showing MCUR1 expression in control fibroblasts and its absence in patient fibroblasts. The molecular weight of the bands is indicated on the left, and tubulin is used as a loading control, 100 µg protein were loaded onto each lane. Bands of four biological replicates were analyzed by densitometry and depicted as a scatter plot with error bars representing the mean ± standard deviation (SD). Statistical significances were determined using an unpaired t-test: *, p < 0.05; **, p < 0.01; Ctrl, Control.

The consanguineous family background indicated a likely autosomal recessive inheritance pattern and we delineated autozygous regions with a total of 128.1 Mbp on chromosomes 4-7 and 16-19 (**Supplementary Figure 3**) with stretches of 70 homozygous SNPs or more using HomozygosityMapper.^22^ Analysis with MutationTaster2^13^ identified a homozygous nonsense mutation in *MCUR1* [chr6:13,798,886G>A GRCh38.p14 | c.802C>T (rs372193345) NM_001031713.4 | p.(R268*)] (**Figure 1B**). In his family, only the affected patient was carrying this mutation homozygously (**Supplementary Figure 4**). In the gnomAD v4.1.0 database [https://gnomad.broadinstitute.org/variant/6-13798886-G-A, accessed at July 23, 2025] this variant is listed with a minor allele frequency of 0.00002365 and never in homozygous state. To assess the physiological relevance of *MCUR1* mutation, we investigated dermal fibroblasts and muscle from the patient and from matched controls. *MCUR1* mRNA copy numbers were reduced in patient tissue as were the protein levels on Western blot. (**Figures 1D-E)**

### MCUR1 deficiency reduces mitochondrial Ca^2+^ influx without disrupting MCU complex assembly and localization

As MCUR1 has been reported to directly bind to MCU and EMRE to form the active MCU complex (**Supplementary** Figure 5A),^10^ we evaluated the mRNA transcription of the MCU subunits in patient muscle and fibroblasts. RNA sequencing did not detect any deviating mRNA copy numbers for the other MCU subunits except for *MCUR1* mRNA (**Supplementary** Figures 5B-C). These findings were confirmed by RT-qPCR (Figure 2A). Interestingly, the absence of MCUR1 did not affect the synthesis of MCU or EMRE proteins in patient cells (Figure 2B) nor the subcellular location of the MCU *puncta* at the inner mitochondrial membrane (Figure 2C).

**Figure 2.**
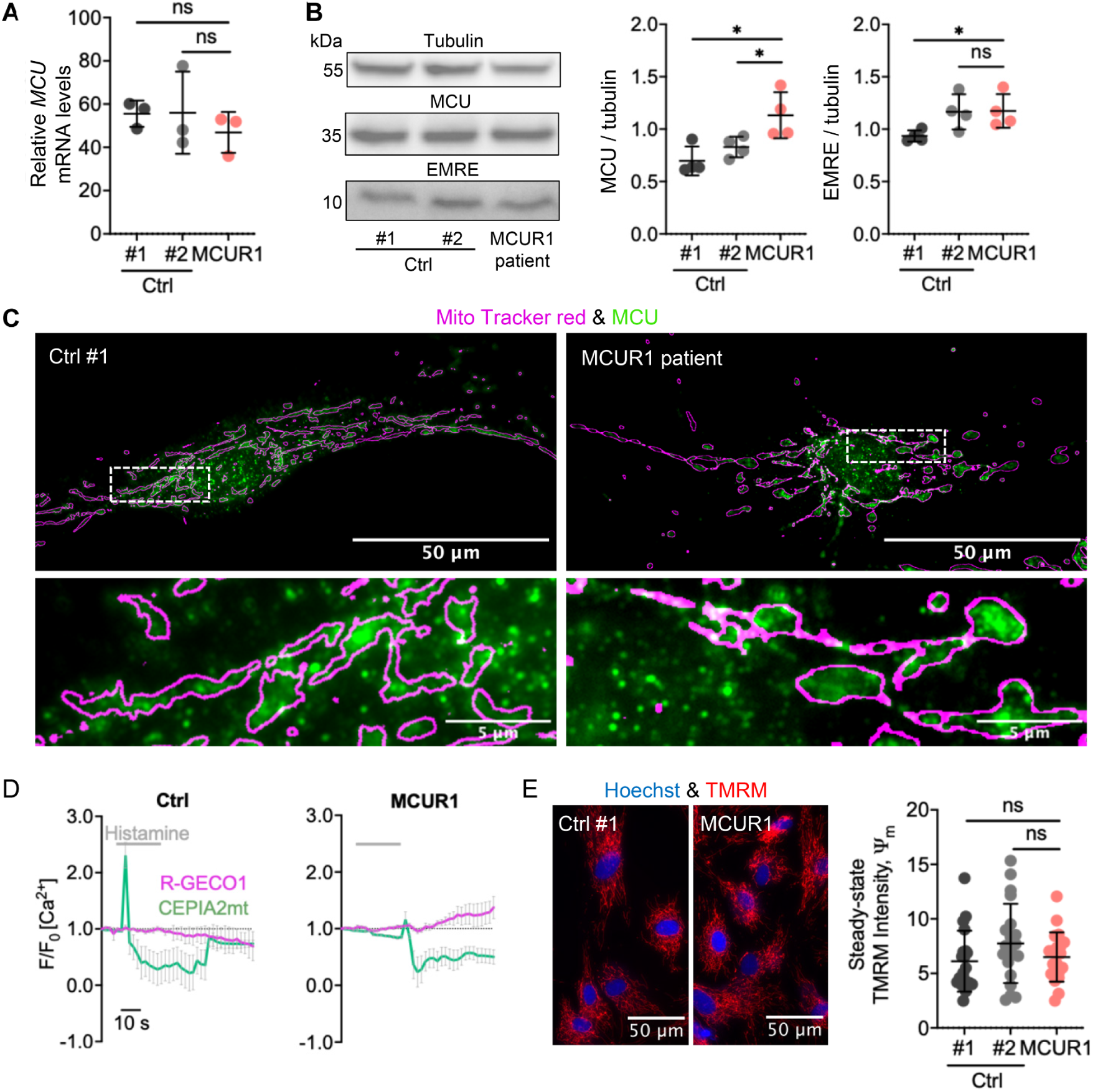
Expression analysis of MCU complex subunits and measurement of mitochondrial Ca^2+^ influx. **(A)** qPCR analysis demonstrating MCU mRNA copy numbers in the MCUR1-deficient patient fibroblasts and controls, error bars represent the mean ± standard deviation (SD) of three biological replicates [*MCU*/1,000 *GAPDH* copies]. **(B)** Immunoblot showing MCU and EMRE protein expression in control and patient fibroblasts. Protein molecular size is indicated on the left; tubulin is used as a loading control, 60 µg protein were loaded onto each lane; error bars represent the mean ± standard deviation (SD) of four biological replicates. **(C)** Immunofluorescence microscopy images showing MCU *puncta* formation (green) in single cells. No difference between patient and controls can be seen. The magenta outlines depict the mitochondrial shapes as generated by staining with MitoTracker Red FM. **(D)** histamine (10 µM) induced cytosolic (magenta) and mitochondrial Ca^2+^ signals (green) were measured using R-GECO1 and CEPIA2mt reporter constructs in primary fibroblasts from the patient compared to controls. Traces show the F/F_0_ ratio of fluorescence in single cells. F_0_ depicts the fluorescence before stimulation. Traces of Ca^2+^ concentrations are shown as mean ± SD from >10 traces of four separate experiments. **(E)** Primary fibroblasts of the patient and from age-matched healthy individuals were stained with TMRM to quantify ψ_m_. Cells were identified by Hoechst 33347 staining of their nuclei. N=20 single cells were analysed in three independent experiments. Scatter blots in A, B, and C show mean ± SD from three separate experiments. For pairwise comparisons t-test were used: *, p < 0.05; ****, p < 0.0001; ns, not significant; Ctrl, control.

To further evaluate the functional consequences of MCUR1 deficiency, we analyzed mtCa^2+^ homeostasis in fibroblasts from our patient and from healthy individuals. Using intraorganellar fluorescent reporter constructs (R-GECO1 and CEPIA2mt),^23^ we measured fluorescence after external stimulation with the Ca^2+^-mobilization agonist histamine at a concentration of 10 μM.

Histamine-induced mtCa^2+^ uptake of the mitochondria was diminished in MCUR1-deficient fibroblasts, while there was a relative increase of the cytosolic Ca^2+^ concentration (Figure 2D) indicating a malfunction of mitochondrial calcium clearing from the cytoplasm after its release from the endoplasmic reticulum (ER).

The major driving force behind the mitochondrial Ca^2+^ uptake is the strongly negative potential across the inner mitochondrial membrane (Ψ_m_), which is maintained by the mitochondrial electron transport chain. To investigate whether a reduction of Ψ_m_ would be present in the patient fibroblast mitochondria as an alternative explanation for the reduced Ca^2+^ influx, we evaluated the Ψ_m_ using the potentiometric fluorescent Tetramethylrhodamine methyl ester (TMRM) dye. However, the *MCUR1* loss-of-function mutation did not alter Ψ_m_, suggesting that MCUR1 mainly regulates Ca^2+^ conductance across the inner mitochondrial membrane (Figure 2E).

### MCUR1 deficiency reduces ATP production and oxygen consumption rate (OCR)

During muscle contraction, Ca^2+^ is released from the sarcoplasmic reticulum into the sarcoplasm, which is then taken up by the mitochondria. The Ca^2+^ influx into the mitochondria regulates the activities of the pyruvate dehydrogenase complex (PDHc), the tricarboxylic acid (TCA) cycle dehydrogenases, the OXPHOS system, and the F_1_F_O_-ATPase thereby coupling ATP demand during muscle work to ATP production.^1,4,5,7^ assess the effect of MCUR1 deficiency on the ATP production, we used a mitochondrial matrix targeted luciferase that reports mitochondrial ATP concentrations *via* light emission. We found a reduced ATP production in the patient fibroblasts

The activities of the isolated OXPHOS complexes and PDHc activity from fibroblasts were normal (**Supplementary** Figure 2). In addition, we performed histochemical analyses of NADH-TR, succinate dehydrogenase (SDH), and cytochrome c oxidase (COX IV) and did not observe any differences compared to controls (**Supplementary** Figure 1B).

Given the central role of mitochondria in energy homeostasis, we investigated whether MCUR1 deficiency affected mitochondrial biogenesis and degradation. Quantification of the mtDNA copy number revealed a significant reduction in the patient-derived fibroblasts suggesting either impaired mitochondrial biogenesis, reduced mtDNA replication, or increased degradation (Figure 3B).

**Figure 3.**
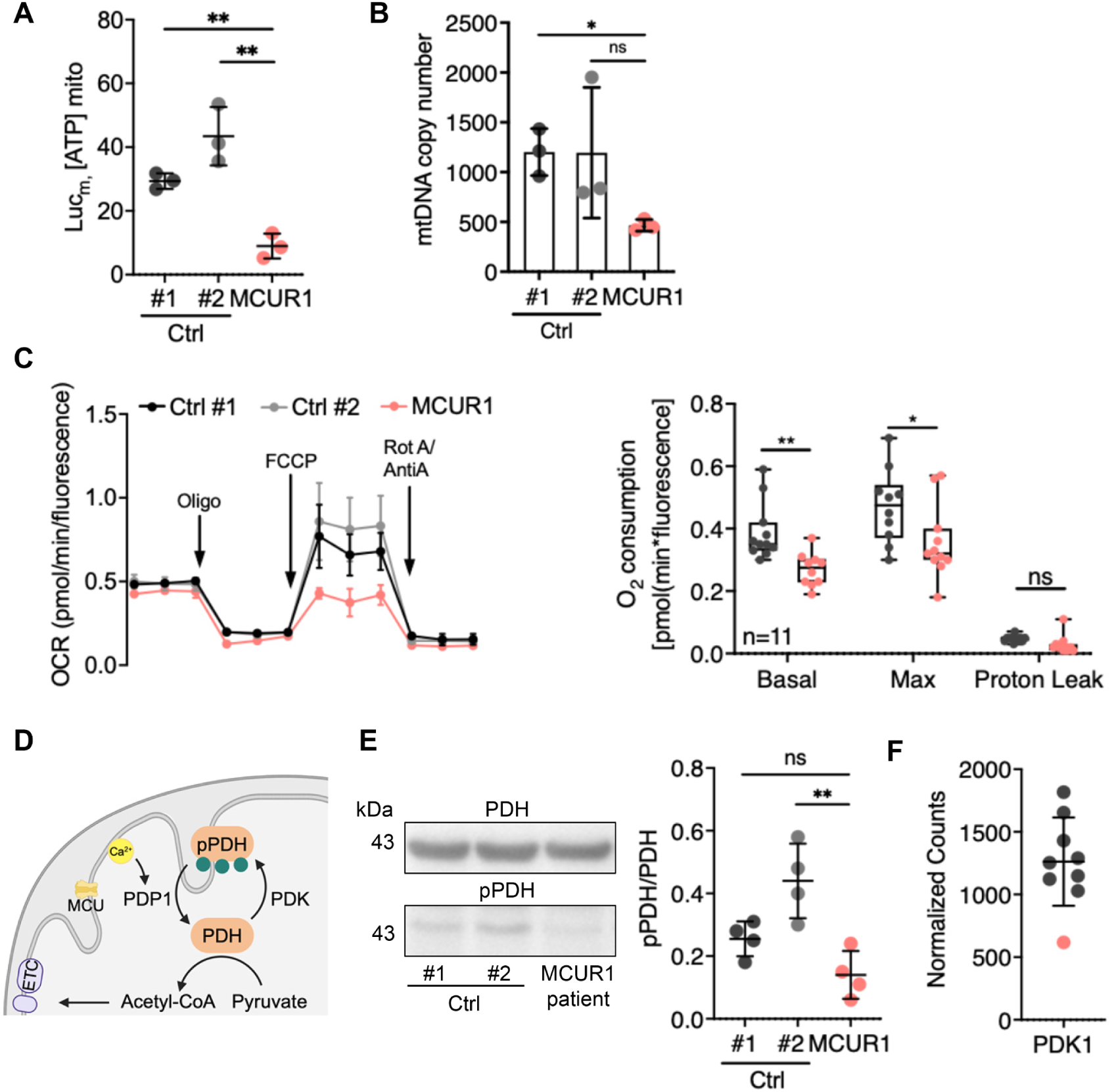
Effect of MCUR1 deficiency on mitochondrial oxygen consumption and mitochondrial energy metabolism. **(A)** Scatterplot of mitochondrial ATP production in control and MCUR1-patient fibroblasts measured by a luminescence assay. **(B)** qRT–PCR analyses for mtDNA copy numbers were performed in controls and MCUR1-patient cells. Scatter plot shows the mean ± SD from three separate experiments. **(C)** Normalized oxygen consumption rates (OCR) of intact control and patient fibroblasts were measured using a Seahorse® flux analyzer. Basal respiration, maximal respiration (in the presence of 1 μM FCCP), and leakage respiration (in the presence of 2.5 μM oligomycin) are depicted. **(D)** Schematic showing the regulation of the pyruvate dehydrogenase complex (PDH). PDH phosphatase subunit 1 (PDP1) dephosphorylates the serine residues of PDH upon activation by Ca^2+^. The graphic is adapted from Wolf *et al.,* 2022^24^ with Biorender. **(E)** Immunoblots from whole cell lysates for quantification of PDH phosphorylation (pPDH, serine 293), normalized to total PDH levels revealed decreased phosphorylation levels of PDH in MCUR1-patient fibroblasts, 100 µg protein were loaded onto each lane. Bands of four biological replicates were analyzed by densitometry and depicted as a scatter plot with error bars representing the mean ± standard deviation (SD). **(F)** Gene expression of PDK1 as normalized counts (by DESeq2). Plot compares controls (black) and MCUR1-patient fibroblasts (red). Statistical significance was determined using an unpaired t-test: **, p < 0.01; *, p < 0.05; ns, not significant; Ctrl, Control.

Although isolated OXPHOS complexes in muscle and the PDHc displayed normal enzymatic activities, we assessed the mitochondrial respiration in intact cells using Seahorse® extracellular flux analysis to assess OXPHOS regulation within the cellular context. MCUR1-deficient fibroblasts exhibited reduced basal oxygen consumption rates (OCR) and a lower maximal respiratory capacity, whereas the proton leak was not affected (Figure 3C).

Given the reduced mtCa^2+^ influx, ATP production and OCR in MCUR1-deficient cells, we hypothesized that PDHc phosphorylation and activity might be altered as seen in elderly sarcopenic patients.^25^ The PDHc can be inactivated by the PDH kinase (PDK) through phosphorylation of three serine residues of the E1-subunit, while activation is achieved by the phospho-PDH phosphatase (PDP1), which is dependent on mtCa^2+^ concentration (Figure 3D). Therefore, we determined the state of phosphorylation of PDHc by immunoblotting. Unexpectedly, this experiment revealed significantly decreased phosphorylation at serine 293 in patient fibroblasts as compared to controls (Figure 3E), suggesting increased PDH enzymatic activity. Consistently, we observed a downregulation of PDK mRNA expression (Figure 3F), indicating that MCUR1-deficient cells may suppress PDK mRNA synthesis in compensation to maintain PDH activity and support mitochondrial metabolism. Interestingly, inhibition of PDK by dichloroacetate (DCA) in elderly mice improved their exercise capacity.^25^(Figure 3A).

### MCUR1 deficiency promotes autophagy pathways in muscle

One potential adaptive response to mitochondrial dysfunction is autophagy.^25^ However, upon serum starvation, the LC3B-II/I ratio increased only slightly without differences between control and patient fibroblast lines. Treatment with an autophagy inhibitor, Bafilomycin A1, caused a strong LC3B-II/I accumulation in all fibroblast lines, indicating intact autophagic flux, but no alteration due to MCUR1 deficiency (**Supplementary** Figure 6). This prompted us to shift our focus to muscle tissue, the primarily affected tissue. Electron microscopy of patient-derived quadriceps muscle revealed swollen mitochondria with disrupted cristae, suggesting mitochondrial damage. Histology revealed large vacuolar structures, suggesting accumulation of autophagic material (Figure 4A). The membranes of these vacuoles were positive for acetylcholine esterase (AChE) activity. In addition, staining for the autophagy-related proteins LAMP2 and LC3B showed abnormal aggregations in the subsarcolemmal regions of the muscle (Figure 4B).

**Figure 4.**
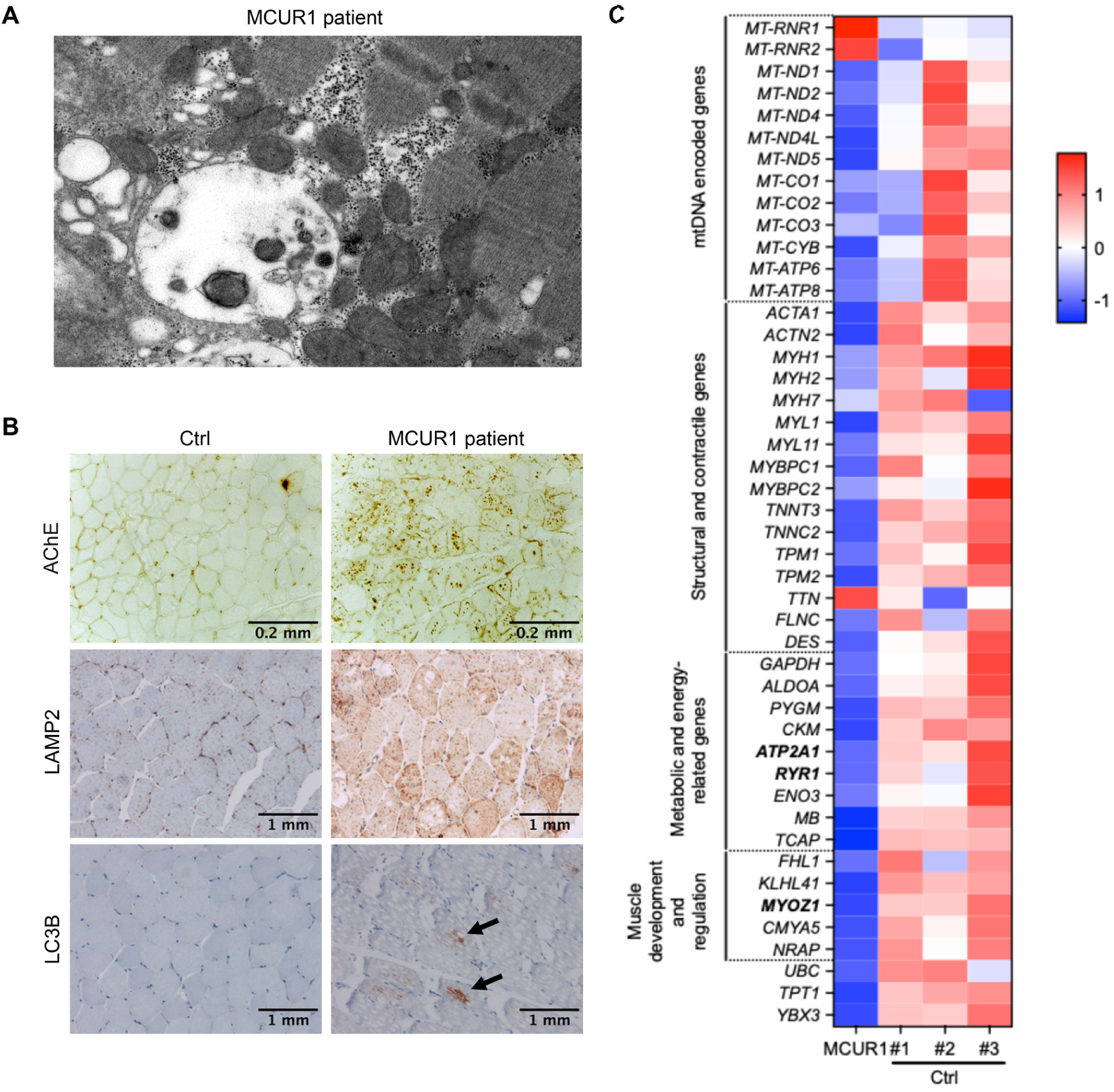
Expression of autophagic markers and RNA-seq analysis of muscle samples. **(A)** Electron microscopy of the muscle of the MCUR1-patient showing an autophagic vacuole. **(B)** Histochemistry of skeletal muscles for acetylcholine esterase activity (AChE), LAMP2, and LC3B. **(C)** Visualization of RNA-seq results with a heatmap suggesting transcriptional changes due to MCUR1 deficiency of muscle tissue from three healthy controls and the MCUR1-patient. Differentially expressed genes (DEGs) are represented with log2 fold changes of ≥0.5, ≤-0.5, and a false discovery rate (FDR) of < 0.05. For pairwise comparisons the t-test was used: *, p < 0.05; ns, not significant; Ctrl, Control.

In order to get a wider perspective of the MCUR1-dependent transcriptional changes, we performed bulk RNA sequencing on muscle samples of three healthy controls *versus* the MCUR1-deficient patient. We found 1992 differentially expressed genes (DEGs), among them, 776 up-regulated and 1216 down-regulated (FDR <0.05; >2 fold change in both directions).

MCUR1 deficiency had caused a clear reduction of the steady state levels of multiple mRNAs that are transcribed from the mtDNA and code for most of the mtDNA coded subunits of the OXPHOS complexes I, II, IV and V. This is in agreement with the reduced mtDNA copy number and could explain the overall reduced OCR and ATP-production. Interestingly, only the mitochondrial ribosomal RNA was highly upregulated, probably as a compensatory measure of the MCUR1-deficient cells. The mRNA of key calcium-handling genes (*RYR1, ATP2A1*) and metabolic enzymes (*CKM, PYGM*) showed reduced mRNA expression. Additionally, we found the broad downregulation of muscle-specific genes (Figure 4C).

## Discussion

In this study, we present the first data on the physiological effects of a biallelic *MCUR1* mutation. We identified a homozygous nonsense variant in *MCUR1* that is associated with vacuolar myopathy. The patient’s leading symptoms include early-onset proximal muscle weakness, generalized muscle atrophy, and cavus foot deformity.

Despite its known role in the MCU complex, we observed that MCUR1 deficiency does not disrupt the assembly or localization of the MCU complex. Expression levels of the channel (MCU, and EMRE) and other regulatory subunits (MICU1, MICU2, and MICU3) were unaltered.^26^ However, MCUR deficiency reduced mtCa^2+^ uptake and increase cytosolic Ca^2+^ accumulation upon histamine stimulation, suggesting a regulatory rather than structural role for MCUR1. Notably, these defects occurred independently of changes in Ψ_m_, supporting the idea that MCUR1 primarily regulates mtCa^2+^ influx. This aligns with prior reports indicating that MCUR1 functions as a critical modulator of mtCa^2+^ influx rather than a core component of the uniporter complex.^10–12^

We observed a reduction of mitochondrial ATP production and of basal oxygen consumption despite preserved enzymatic activities of the PDH and OXPHOS complexes. This metabolic impairment may, at least in part, be attributed to a reduced mtDNA copy number.

In agreement with previous studies demonstrating that modulation of MCU or MCUR1 in skeletal muscle regulates oxidative metabolism, mitochondrial ATP output, and overall muscle function,^7,26,27^ we show that the *MCUR1* mutation disrupts the cascade of mtCa^2+^ influx, ATP production, and ultimately respiratory energy production in skeletal muscle. The PDHc is activated *via* dephosphorylation by the Ca^2+^-dependent PDP, thereby coupling anaerobic glycolysis with aerobic ATP production through the TCA cycle and OXPHOS. In aging muscle, PDHc activity declines though a mechanism that seems to involve MCUR1.^26^ Inhibition of the PDK by DCA was able to restore PDHc activity and overcome the bioenergetic defects.^26^ Similarly, the intrinsic downregulation of PDK observed in the MCUR1-deficient patient may represent a compensatory response to sustain PDHc activity and meet muscular ATP demand despite impaired Ca^2+^-dependent metabolic regulation.

Knockout of *MCUR1* or its activity loss has been linked to reduced muscle mass and strength in mouse models and *MCUR1* was found to be downregulated on the mRNA level in older sarcopenic patients, confirming that MCUR1 has a role in muscle regeneration.^10–12,25,26^

Muscle biopsy samples of our patient revealed the presence of autophagic vacuoles with sarcolemmal features (AVSF). These are distinctive findings of autophagic vacuolar myopathies (AVMs) such as Danon disease and X-linked myopathy with excessive autophagy (XMEA). These diseases are characterized by intracytoplasmic vacuoles that express sarcolemmal proteins on their surface and exhibit acetylcholinesterase activity. This characteristic staining pattern effectively distinguishes AVSF myopathies from other vacuolar myopathies, directing clinicians toward the testing of *LAMP2* for Danon disease or *VMA21* for XMEA. LAMP-2 deficiency impairs autophagosome-lysosome fusion and chaperone-mediated autophagy, while VMA21 deficiency disrupts the lysosomal V-ATPase assembly, elevating lysosomal pH and decreasing their degradative capacity.^21^ Autophagosome-lysosome fusion, a critical step in autophagy, which requires ATP for multiple processes, including proton pumping by V-ATPases to maintain lysosomal acidity and soluble N-ethylmaleimide-sensitive-factor attachment receptor (SNARE)-mediated vesicle fusion.^25,28^

When MCUR1 dysfunction impairs mitochondrial calcium uptake, muscle mitochondria lose their ability to sequester calcium during physiological signaling processes. Sarcoplasmic calcium clearance becomes dependent on slower, low-capacity mechanisms including SERCA pumps and plasma membrane exchangers. This creates both spatial and temporal dysregulation of the entire contractile apparatus of the muscle.

In summary, MCUR1 deficiency and subsequent mitochondrial Ca^2+^ uptake impairment, appears to create a complex cellular phenotype leading to diminished mitochondrial function despite the functional integrity of all its players, ultimately leading to ATP deficiency and deranged autophagy. A complete picture, however, would require the investigation of a larger patient cohort. These findings expand the understanding of AVSF-related myopathies by identifying MCUR1 deficiency as a new pathogenic principle.

## Data Availability

Patient genomic (DNA) and transcriptomic (RNA) data cannot be freely submitted to a repository. The novel variant has been submitted to ClinVar (SCV006307813). Genetic data are available from the authors upon reasonable request within the frame of the patient’s consent.

Raw data were generated at Charité-Universitätsmedizin Berlin, Corporate Member of Freie Universität Berlin, Humboldt-Universität zu Berlin, and Berlin Institute of Health (BIH), Department of Neuropediatrics, Berlin, Germany. The RNAseq dataset generated in this study is available from the corresponding author upon reasonable request. The novel variant was submitted to the ClinVar database (https://www.ncbi.nlm.nih.gov/clinvar; accession number SCV006307813).

## Acknowledgements

We thank the patient and his family for their participation in this study. We thank Richard Rodenburg, Nijmegen Center for Mitochondrial Disorders, Netherlands for measuring the OXPHOS activities in patient fibroblasts.

## Supplementary Figures

**Supplementary Figure 1.**
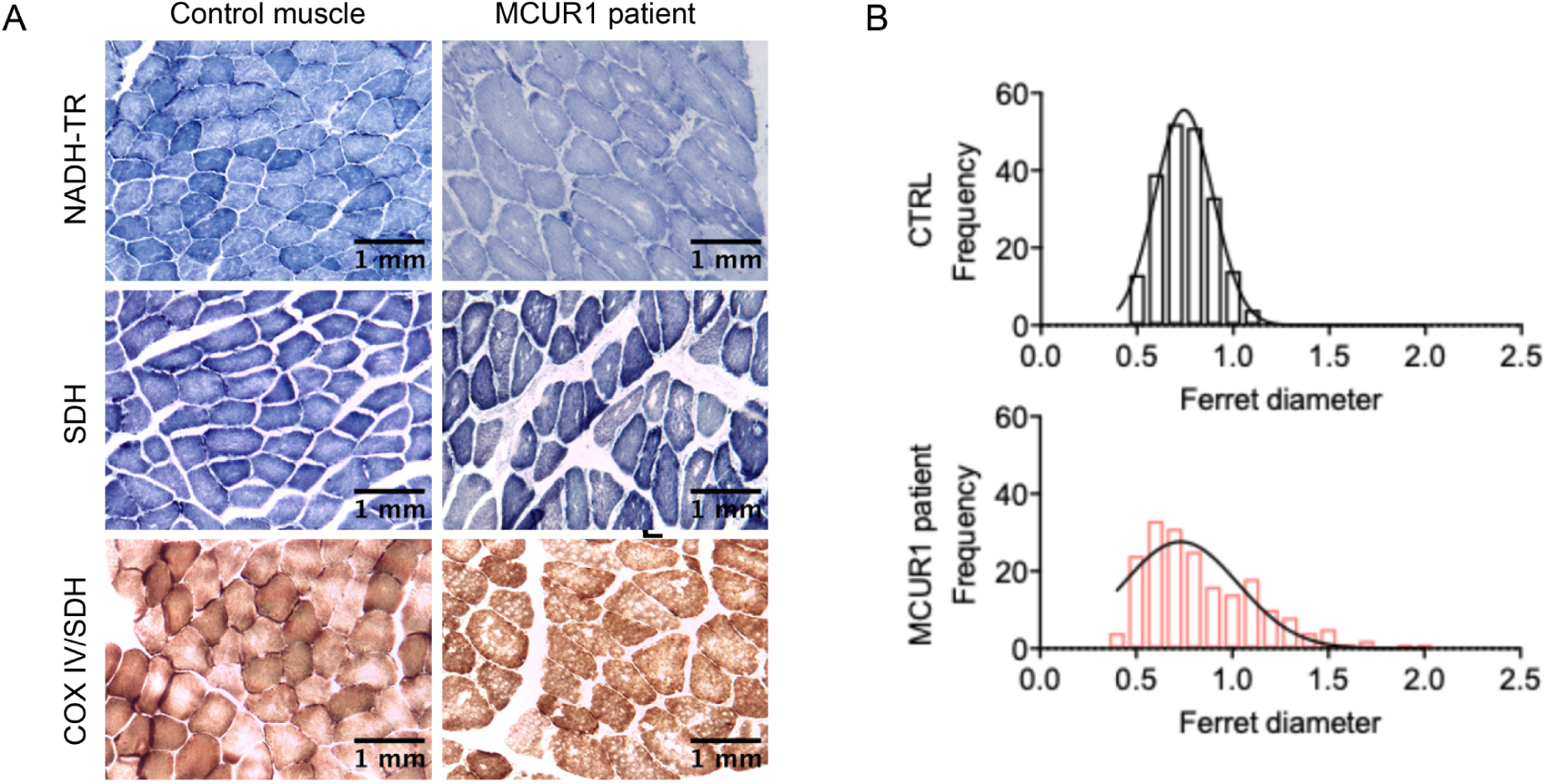
**(A)** Mitochondria specific histologic stainings of NADH-TR (complex I), SDH (Complex II), and COX (complex IV) show normal staining patterns, the patient’s muscle shows multiple intrafiber vacuolar deposits. **(B)** Mean fiber size and size dispersion of healthy control and MCUR1-patient muscle (approx. 200 fibers per muscle; n = 6 sections analyzed); the patient’s muscle shows a wider dispersion of muscle fiber sizes with a slight shift of the mean fiber diameter to the left.

**Supplementary Figure 2.**
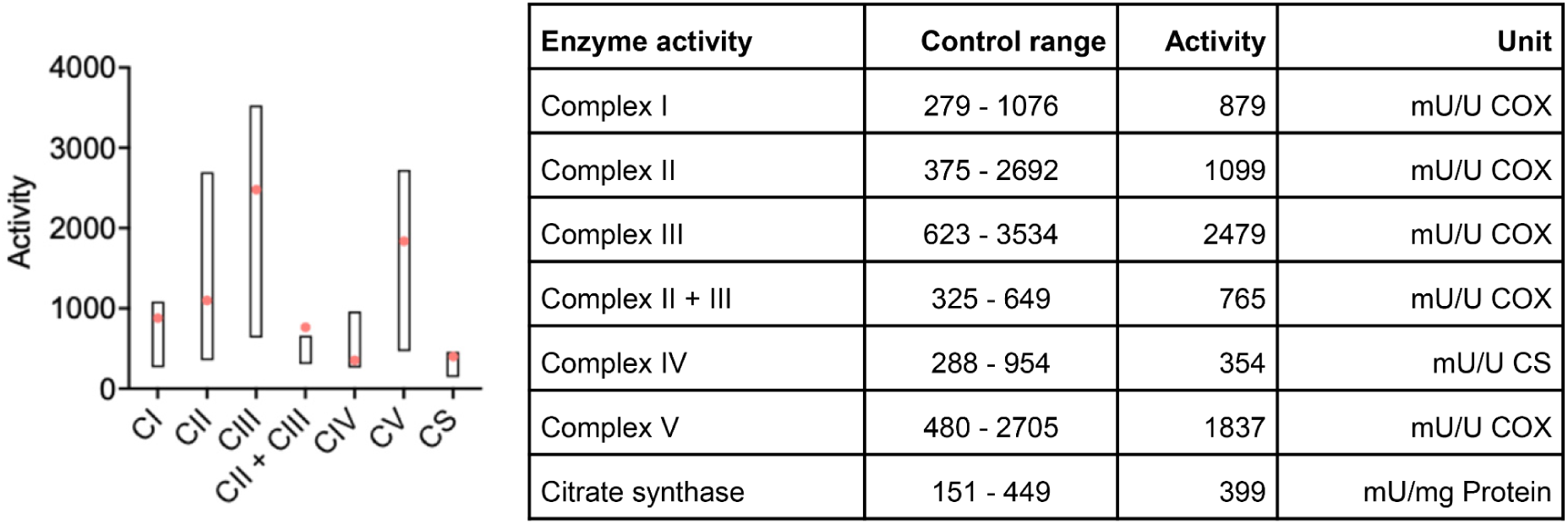
Respiratory chain complex activities in cultured skin fibroblasts of the patient (indicated in red) in comparison to reference values.

**Supplementary Figure 3.**
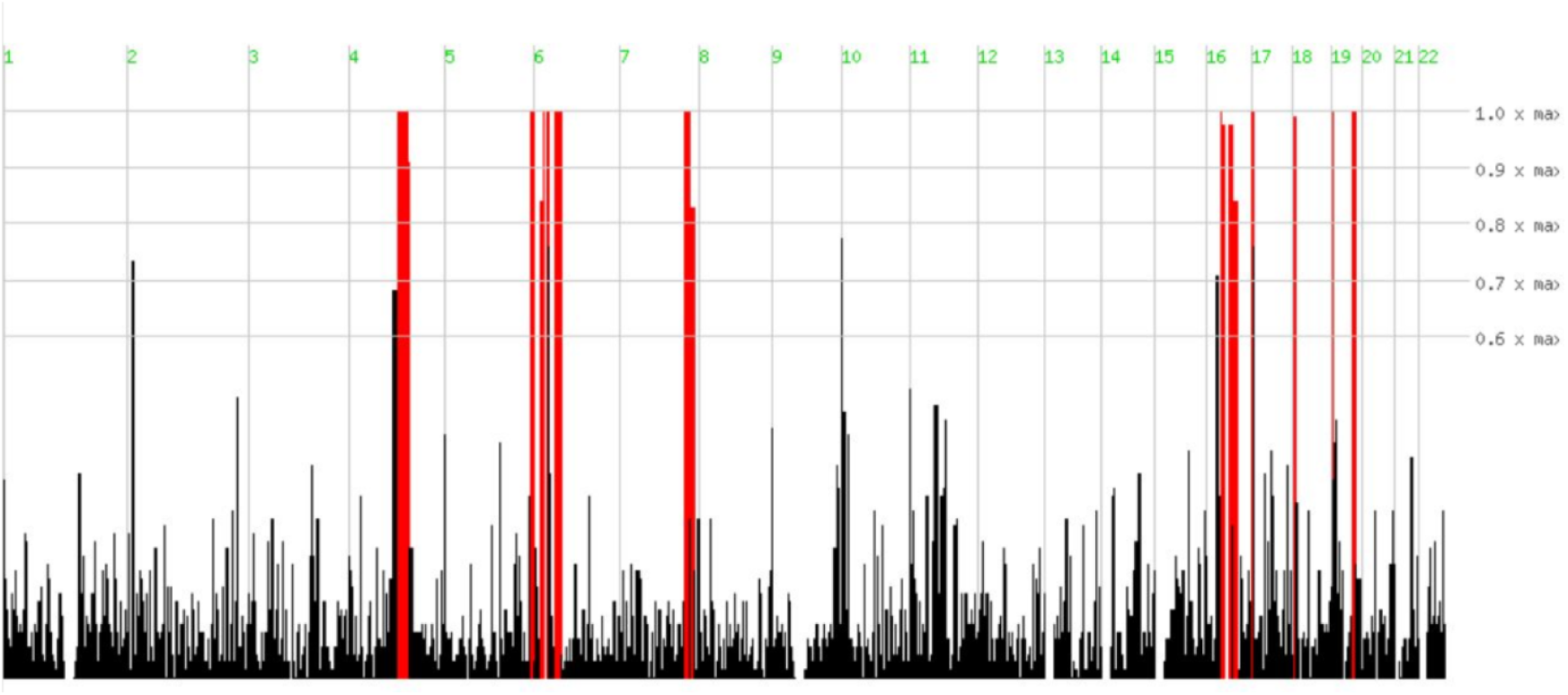
Regions that are autozygous for >70 SNPs in the exome of the index patient as delineated by the Homozygositymapper software. The entire autozygous region stretches over 128.1 Mbp. The *MCUR1* gene is located at chr6:13,786,781-13,814,792 (GRCh37).

**Supplementary Figure 4.**
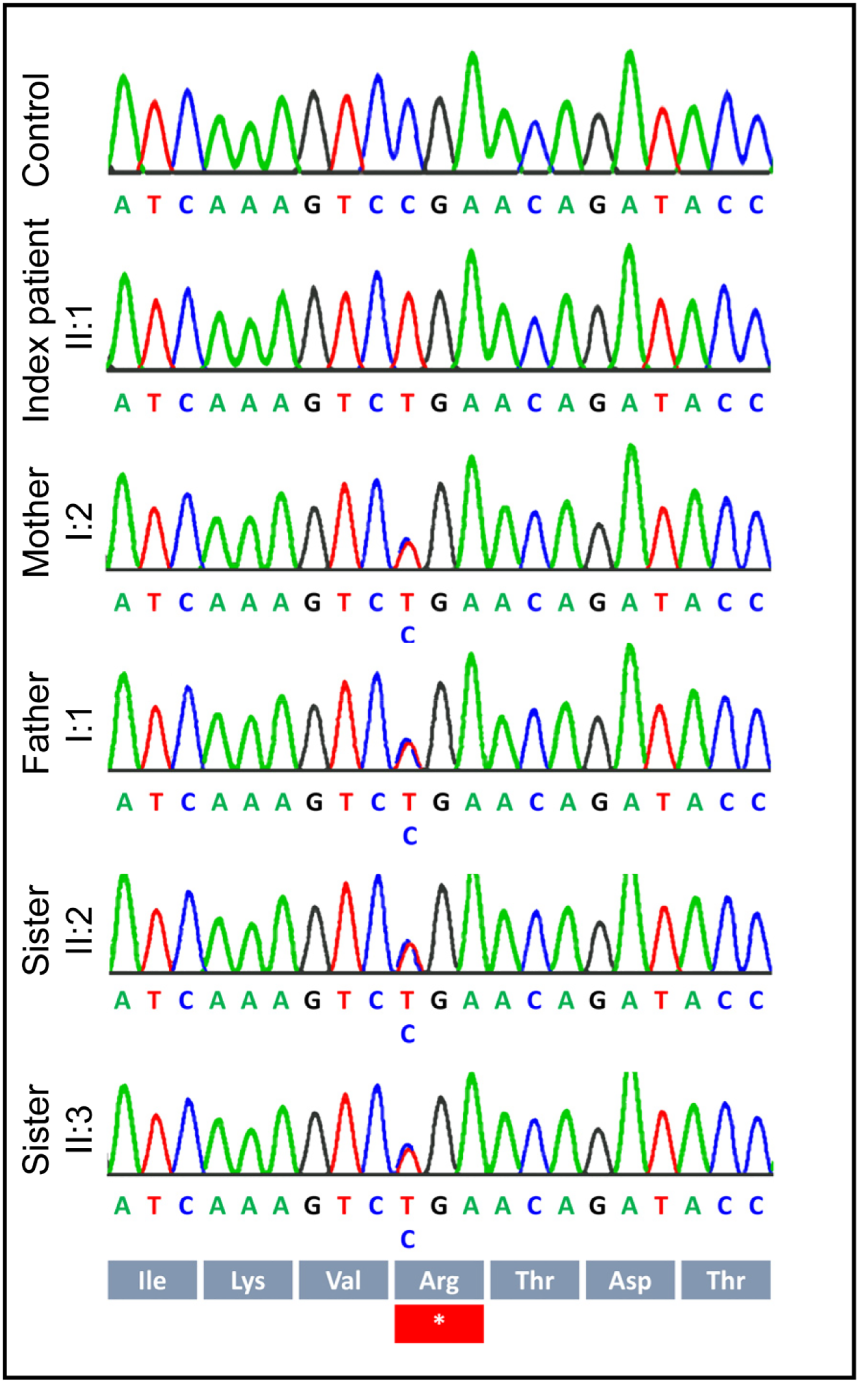
Segregation of the *MCUR1* variant [c.802C>T, NM_001031713.4 | p.(R268*)] in the patient’s family.

**Supplementary Figure 5.**
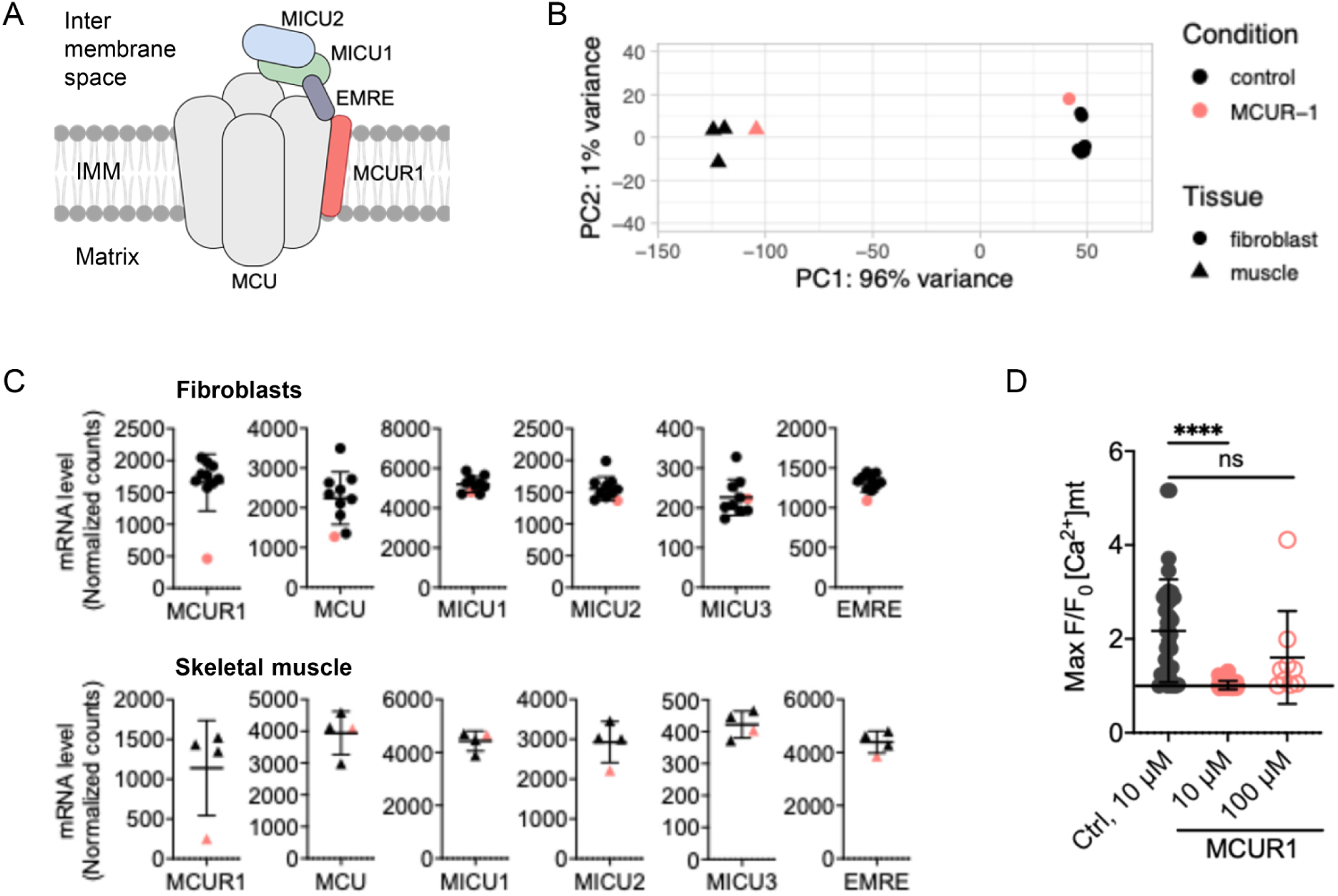
**(A)** Schematic overview of the MCU complex and its protein subunits. **(B)** Principal component analysis (PCA) distinguishes between fibroblasts (patient and n=3 controls) and muscle samples (patient and n=3 controls). **(C)** mRNA expression of genes of the MCU complex measured by RNA sequencing of fibroblasts (n = 9 age-matched controls *versus* the patient) and human muscle biopsy samples (n = 3 age-matched controls versus the patient). **(D)** Histamine-induced (10 µM) mitochondrial Ca^2+^ signals from the affected individual compared to controls. The scatter plot demonstrates the peak increase of mtCa^2+^ concentrations against the basic steady state concentrations [F/F_0_ after histamine (10 µM and 100 µM) treatment. Traces of mtCa^2+^ concentrations are shown as mean ± SD from four separate experiments.

**Supplementary Figure 6.**
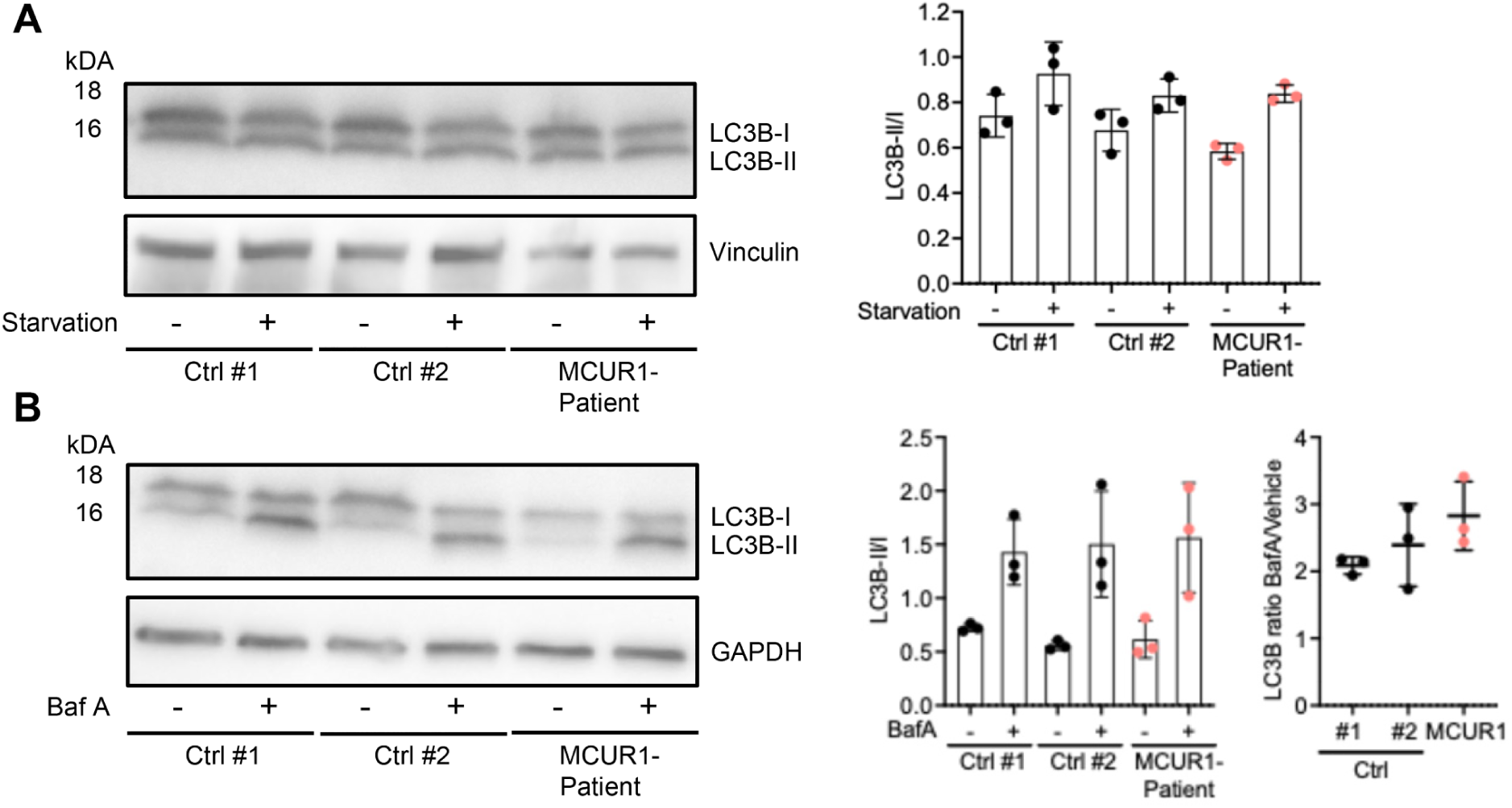
Quantification of LC3 levels in the presence or absence of bafilomycin A1 in starved and fed cultured dermal fibroblasts. **(A)** Western blot of LC3 protein levels in fibroblasts that had been starved by serum deprivation (+) and unstarved controls (-). Band densities were analyzed by densitometry and LC3B-II levels were normalized to LC3B-I. **(B)** Autophagic flux measured *via* theLC3B-II/I ratio after inhibition with bafilomycin A in starved control and patient fibroblasts cells. No difference was seen between control and patient lines, indicating normal autophagy in fibroblasts.

## Supplemental Tables

**Table 1:**
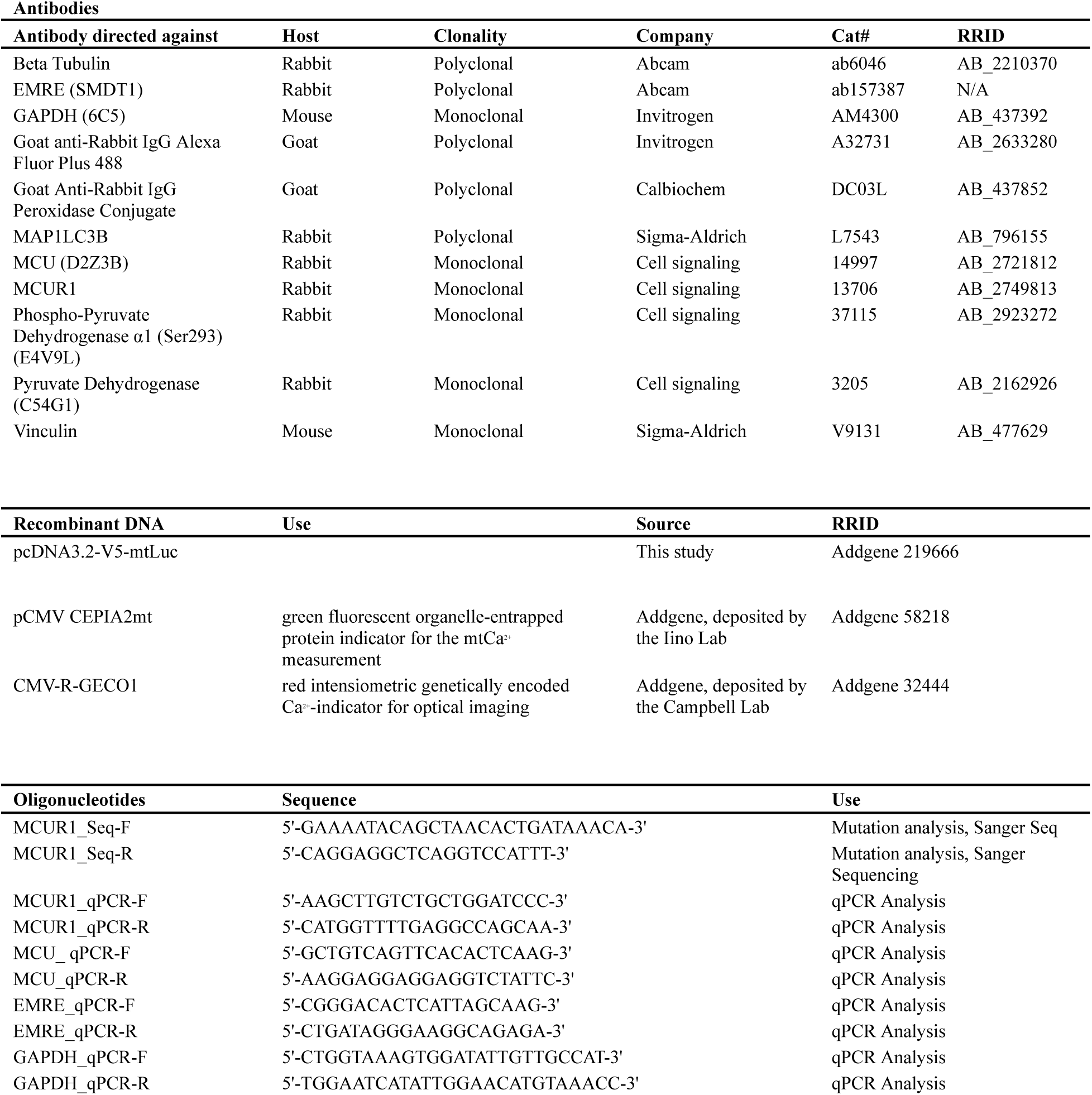
Materials used in this study.

## References

1. Giorgi C, Marchi S, Pinton P. The machineries, regulation and cellular functions of mitochondrial calcium. Nat Rev Mol Cell Biol. 2018;19(11):713–730. doi:10.1038/s41580-018-0052-8

2. Baughman JM, Perocchi F, Girgis HS, et al. Integrative genomics identifies MCU as an essential component of the mitochondrial calcium uniporter. Nature. 2011;476(7360):341–345. doi:10.1038/nature10234

3. De Stefani D, Raffaello A, Teardo E, Szabò I, Rizzuto R. A forty-kilodalton protein of the inner membrane is the mitochondrial calcium uniporter. Nature. 2011;476(7360):336–340. doi:10.1038/nature10230

4. De Stefani D, Rizzuto R. Molecular control of mitochondrial calcium uptake. Biochem Biophys Res Commun. 2014;449(4):373–376. doi:10.1016/j.bbrc.2014.04.142

5. Pan X, Liu J, Nguyen T, et al. The physiological role of mitochondrial calcium revealed by mice lacking the mitochondrial calcium uniporter. Nat Cell Biol. 2013;15(12):1464–1472. doi:10.1038/ncb2868

6. Chemello F, Mammucari C, Gherardi G, Rizzuto R, Lanfranchi G, Cagnin S. Gene expression changes of single skeletal muscle fibers in response to modulation of the mitochondrial calcium uniporter (MCU). Genomics Data. 2015;5:64–67. doi:10.1016/j.gdata.2015.05.023

7. Mammucari C, Gherardi G, Zamparo I, et al. The Mitochondrial Calcium Uniporter Controls Skeletal Muscle Trophism In Vivo. Cell Rep. 2015;10(8):1269–1279. doi:10.1016/j.celrep.2015.01.056

8. Logan CV, Szabadkai G, Sharpe JA, et al. Loss-of-function mutations in MICU1 cause a brain and muscle disorder linked to primary alterations in mitochondrial calcium signaling. Nat Genet. 2014;46(2):188–193. doi:10.1038/ng.2851

9. Debattisti V, Horn A, Singh R, et al. Dysregulation of Mitochondrial Ca2+ Uptake and Sarcolemma Repair Underlie Muscle Weakness and Wasting in Patients and Mice Lacking MICU1. Cell Rep. 2019;29(5):1274–1286.e6. doi:10.1016/j.celrep.2019.09.063

10. Tomar D, Dong Z, Shanmughapriya S, et al. MCUR1 Is a Scaffold Factor for the MCU Complex Function and Promotes Mitochondrial Bioenergetics. Cell Rep. 2016;15(8):1673–1685. doi:10.1016/j.celrep.2016.04.050

11. Mallilankaraman K, Cárdenas C, Doonan PJ, et al. MCUR1 is an essential component of mitochondrial Ca2+ uptake that regulates cellular metabolism. Nat Cell Biol. 2012;14(12):1336–1343. doi:10.1038/ncb2622

12. Vais H, Tanis JE, Müller M, Payne R, Mallilankaraman K, Foskett JK. MCUR1, CCDC90A, Is a Regulator of the Mitochondrial Calcium Uniporter. Cell Metab. 2015;22(4):533–535. doi:10.1016/j.cmet.2015.09.015

13. Schwarz JM, Cooper DN, Schuelke M, Seelow D. MutationTaster2: mutation prediction for the deep-sequencing age. Nat Methods. 2014;11(4):361–362. doi:10.1038/nmeth.2890

14. Ewels P, Magnusson M, Lundin S, Käller M. MultiQC: summarize analysis results for multiple tools and samples in a single report. Bioinformatics. 2016;32(19):3047–3048. doi:10.1093/bioinformatics/btw354

15. Dobin A, Davis CA, Schlesinger F, et al. STAR: ultrafast universal RNA-seq aligner. Bioinformatics. 2013;29(1):15–21. doi:10.1093/bioinformatics/bts635

16. Li H, Handsaker B, Wysoker A, et al. The Sequence Alignment/Map format and SAMtools. Bioinformatics. 2009;25(16):2078–2079. doi:10.1093/bioinformatics/btp352

17. Pertea M, Pertea GM, Antonescu CM, Chang TC, Mendell JT, Salzberg SL. StringTie enables improved reconstruction of a transcriptome from RNA-seq reads. Nat Biotechnol. 2015;33(3):290–295. doi:10.1038/nbt.3122

18. Zhu A, Ibrahim JG, Love MI. Heavy-tailed prior distributions for sequence count data: removing the noise and preserving large differences. Bioinformatics. 2019;35(12):2084–2092. doi:10.1093/bioinformatics/bty895

19. Ge SX, Jung D, Yao R. ShinyGO: a graphical gene-set enrichment tool for animals and plants. Bioinformatics. 2020;36(8):2628–2629. doi:10.1093/bioinformatics/btz931

20. Zink A, Haferkamp U, Wittich A, Beller M, Pless O, Prigione A. High-content screening of mitochondrial polarization in neural cells derived from human pluripotent stem cells. STAR Protoc. 2022;3(3):101602. doi:10.1016/j.xpro.2022.101602

21. Stenzel W, Nishino I, von Moers A, et al. Juvenile autophagic vacuolar myopathy – a new entity or variant? Neuropathol Appl Neurobiol. 2013;39(4):449–453. doi:10.1111/nan.12018

22. Seelow D, Schuelke M. HomozygosityMapper2012--bridging the gap between homozygosity mapping and deep sequencing. Nucleic Acids Res. 2012;40(W1):W516–W520. doi:10.1093/nar/gks487

23. Suzuki J, Kanemaru K, Ishii K, Ohkura M, Okubo Y, Iino M. Imaging intraorganellar Ca2+ at subcellular resolution using CEPIA. Nat Commun. 2014;5:4153. doi:10.1038/ncomms5153

24. Wolf C, Pouya A, Bitar S, et al. GDAP1 loss of function inhibits the mitochondrial pyruvate dehydrogenase complex by altering the actin cytoskeleton. Commun Biol. 2022;5(1):541. doi:10.1038/s42003-022-03487-6

25. Nichenko AS, Southern WM, Qualls AE, et al. Mitochondrial dysfunction and autophagy responses to skeletal muscle stress. bioRxiv. Preprint posted online April 4, 2019:597476. doi:10.1101/597476

26. Gherardi G, Weiser A, Bermont F, et al. Mitochondrial calcium uptake declines during aging and is directly activated by oleuropein to boost energy metabolism and skeletal muscle performance. Cell Metab. 2025;37(2):477–495.e11. doi:10.1016/j.cmet.2024.10.021

27. Gherardi G, Nogara L, Ciciliot S, et al. Loss of mitochondrial calcium uniporter rewires skeletal muscle metabolism and substrate preference. Cell Death Differ. 2019;26(2):362–381. doi:10.1038/s41418-018-0191-7

28. Bootman MD, Chehab T, Bultynck G, Parys JB, Rietdorf K. The regulation of autophagy by calcium signals: Do we have a consensus? Cell Calcium. 2018;70:32–46. doi:10.1016/j.ceca.2017.08.005

